# The accumulation of erythrocytes quantified and visualized by Glycophorin C in carotid atherosclerotic plaque reflects intraplaque hemorrhage and pre-procedural neurological symptoms

**DOI:** 10.1101/2021.07.15.21260570

**Authors:** Joost M. Mekke, Tim R. Sakkers, Maarten C. Verwer, Noortje A.M. van den Dungen, Y. Song, C. Miller, Aloke V Finn, Gerard Pasterkamp, Michal Mokry, Hester M. den Ruijter, Aryan Vink, Dominique P. V. de Kleijn, Gert J. de Borst, Saskia Haitjema, Sander W. van der Laan

## Abstract

**Background and aims:** The accumulation of erythrocyte membranes within an atherosclerotic plaque may contribute to the deposition of free cholesterol and thereby the enlargement of the necrotic core. Erythrocyte membranes can be visualized and quantified in the plaque by immunostaining for the erythrocyte marker glycophorin C. Consequently, we hypothesized, that the accumulation of erythrocytes quantified by glycophorin C is a marker for plaque vulnerability and may therefore reflect intraplaque hemorrhage (IPH), vulnerability of plaques and predict pre-procedural neurological symptoms.

**Methods:** We employed the `CellProfiler`-integrated `slideToolKit` workflow to visualize and quantify glycophorin C, defined as the total plaque area that is positive for glycophorin C, in single slides of culprit lesions obtained from the Athero-Express Biobank of 1,819 consecutive asymptomatic and symptomatic patients who underwent carotid endarterectomy. Our assessment included the evaluation of various parameters such as lipid core, calcifications, collagen content, SMC content, and macrophage burden. These parameters were evaluated using a semi-quantitative scoring method, and the resulting data was dichotomized as predefined criteria into categories of no/minor or moderate/heavy staining. In addition, the presence or absence of IPH was also scored.

**Results:** The prevalence of IPH and pre-procedural neurological symptoms were 62.4% and 87.1%, respectively. The amount of glycophorin staining was significantly higher in samples from men compared to samples of women (median 7.15 (IQR:3.37, 13.41) versus median 4.06 (IQR:1.98, 8.32), p<0.001). Glycophorin C was associated with IPH adjusted for clinical confounders (OR 1.90; 95% CI 1.63, 2.21; p=<0.001). Glycophorin C was significantly associated with ipsilateral pre-procedural neurological symptoms (OR:1.27, 95%CI:1.06-1.41, *p*=0.005). Sex-stratified analysis, showed that this was also the case for men (OR 1.37; 95%CI 1.12, 1.69; p=0.003), but not for women (OR 1.15; 95%CI 0.77, 1.73; p=0.27). Glycophorin C was associated with classical features of a vulnerable plaque, such as a larger lipid core, a higher macrophage burden, less calcifications, a lower collagen and SMC content. There were marked sex differences, in men, glycophorin C was associated with calcifications and collagen while these associations were not found in women.

**Conclusions:** The accumulation of erythrocytes in atherosclerotic plaque quantified and visualized by glycophorin C was independently associated with the presence of IPH, preprocedural symptoms in men, and with a more vulnerable plaque composition in both men and women. These results strengthen the notion that the accumulation of erythrocytes quantified by glycophorin C can be used as a marker for plaque vulnerability.

## Introduction

Atherosclerotic plaque features such as, a large lipid core, a thin fibrous cap, a large infiltrate of pro-inflammatory cells, neovascularization and intraplaque hemorrhage (IPH) contribute to plaque instability^1,2^. These features are a result of processes contributing to the conversion of stable, asymptomatic lesion to an unstable, rupture prone or ruptured plaque, of which inflammation, cellular apoptosis and expansion of the lipid-rich necrotic core are the most known and studied^2^.

Macrophage and smooth-muscle foams cell death has been recognized as major contributor to the expansion of the lipid^3^. Additionally, IPH was identified as a crucial contributor to the accumulation of extracellular free cholesterol, and thus the expansion of the lipid core, within unstable plaques. This is thought to be due to the accumulation of extravasated erythrocytes, which are a large component of IPHs^4^. Because of the high cholesterol content in the membranes of erythrocytes, which is higher than that of all other cells in the body, with lipids constituting 40% of their weight, erythrocytes can supply a significant amount of lipids to the necrotic core^5^. The accumulation of intact erythrocytes and erythrocytes membranes within atherosclerotic plaque, and thus recent and older plaque hemorrhages, can be visualized by immunostaining against glycophorins^4^. Glycophorins are erythrocyte specific membrane proteins that represent about 2% of the erythrocyte membrane proteins, of whom glycophorin A is the most prevalent with 5∼9×10^5^ copies per cell, while glycophorin C is present in lower amounts, with approximately 0.5∼1.0×10^5^ copies in each red blood cell^6^.

Previous smaller studies have used a similar approach to quantify erythrocyte membranes by staining for glycophorin A. However, those studies used either a semi quantitative subjective score, with higher scores indicating higher percentages^4,7^, or a ratio^8^, rather than a continuous scale as in this study. In these studies, higher glycophorin scores, in either carotid^8^ or coronary specimen^4^, were associated with more advanced plaque and with plaque features that reflect plaque unstability such as a large necrotic core, increased neovascularization, and increased macrophage infiltration^4,7^. The accumulation of erythrocyte has not been visualized and quantified by staining for glycophorin C before in a large and harmonized study sample. To accommodate the standardardized and automatic analysis of whole-slide histological images (WSI), and overcome the drawbacks of the inter- and intra-observer variability of manual assessment methods, the `slideToolKit` workflow was developed^9^. The foundation of `slideToolKit` rests upon `CellProfiler`^10^, an image-analysis method that utilizes supervised learning to accurately quantify the total area positive for the glycophorin C stain. By integrating this method into our `slideToolKit`, we have developed a comprehensive automated workflow^9^. Consequently, the utilization of this workflow for WSI analyses offers several compelling advantages over the labor-intensive and semi-quantitative manual assessment method^11,12^. Specifically, it enables increased accuracy, enhanced reproducibility, and reduced requirement for human involvement^13^.

We hypothesized that the accumulation of erythrocytes visualized and quantified by glycophorin C is a marker for plaque vulnerability and that it is associated with IPH and pre-procedural neurological symptoms. Furthermore, in this study, we explored the association of glycophorin C with other binary plaque characteristics, such as lipid core size, macrophage burden, collagen- and smooth muscle cell (SMC) content, to examine its effect on the plaque composition. Given our previous findings, showing IPH is more prevalent in men and that IPH is a predictor of future major adverse cardiovascular events (MACE) only in men, we conducted a separate sex-stratified analysis to clarify and investigate possible sex differences^14^. Finally, we associated glycophorin C with secondary MACE within three-years of follow up.

## Results

### Study population

A total of 1,819 CEA patients from the Athero-Express Biobank were included in the current analysis (mean age 69±9 years, 29.8% women, 87.1% symptomatic presentation, **Table 1**). Most patients had one or more cardiovascular comorbidities such as hypertension (75%), hypercholesterolemia (70%), diabetes (25%), or a history of coronary artery disease (CAD, 33%). Women were more often diagnosed with hypertension, had significantly higher levels of total cholesterol, LDL and HDL, a significantly worse kidney function, were more often smokers and were more likely to be diagnosed with a critical carotid stenosis (70-99%), but less likely to have a history of CAD, in comparison with men (**Table 1**). Moreover, the total plaque area covered by glycophorin C staining adjusted for the plaque size, expressed as percentage glycophorin C staining was significantly higher in men compared to women (**Table 1**).

**Table 1.** Baseline patient Characteristics

| Variable | Overall, N = 1,819<sup>1</sup> | Male, N = 1,277<sup>1</sup> | Female, N = 542<sup>1</sup> | p-value<sup>2</sup> |
| --- | --- | --- | --- | --- |
| **Age - years** | 69 (9) | 69 (9) | 70 (10) | 0.21 |
| **Systolic BP – mmHg** | 153 (26) | 153 (25) | 153 (26) | 0.72 |
| **Diastolic BP – mmHg** | 82 (28) | 82 (25) | 81 (34) | 0.076 |
| **Hypertension** | 1,345/1,787 (75%) | 915/1,255 (73%) | 430/532 (81%) | <0.001 |
| **Diabetes mellitus** | 447/1,819 (25%) | 319/1,277 (25%) | 128/542 (24%) | 0.54 |
| **Hypercholesterolemia** | 1,187/1,687 (70%) | 833/1,192 (70%) | 354/495 (72%) | 0.50 |
| **Total cholesterol – mmol/L** | 4.64 (3.90, 5.51) | 4.50 (3.81, 5.34) | 5.00 (4.03, 5.91) | <0.001 |
| **LDL – mmol/L** | 2.68 (2.07, 3.44) | 2.60 (2.02, 3.30) | 2.79 (2.20, 3.78) | 0.002 |
| **HDL – mmol/L** | 1.12 (0.93, 1.40) | 1.07 (0.90, 1.30) | 1.30 (1.05, 1.60) | <0.001 |
| **Triglycerides – mmol/L** | 1.46 (1.07, 2.07) | 1.50 (1.07, 2.10) | 1.40 (1.09, 1.95) | 0.19 |
| **Body Mass Index – kg/m2** | 26.1 (24.1, 28.7) | 26.2 (24.2, 28.7) | 25.7 (23.2, 28.7) | 0.012 |
| **eGFR – mL/min/1.73/m2** | 73.2 (21.5) | 74.5 (21.5) | 70.3 (21.1) | <0.001 |
| **Smoking Status** |  |  |  | <0.001 |
| *Current smoker* | 612/1,731 (35%) | 396/1,222 (32%) | 216/509 (42%) |  |
| *Ex-smoker* | 886/1,731 (51%) | 691/1,222 (57%) | 195/509 (38%) |  |
| *Never smoked* | 233/1,731 (13%) | 135/1,222 (11%) | 98/509 (19%) |  |
| **History of CAD** | 602/1,818 (33%) | 471/1,277 (37%) | 131/541 (24%) | <0.001 |
| **History of PAOD** | 383/1,818 (21%) | 272/1,277 (21%) | 111/541 (21%) | 0.71 |
| **History of Peripheral interventions** | 381/1,812 (21%) | 257/1,272 (20%) | 124/540 (23%) | 0.19 |
| **History of TIA/Stroke** | 1,426/1,819 (78%) | 996/1,277 (78%) | 430/542 (79%) | 0.53 |
| **Use of antihypertensive drugs** | 1,380/1,814 (76%) | 967/1,275 (76%) | 413/539 (77%) | 0.72 |
| **Use of statins** | 1,444/1,813 (80%) | 1,011/1,274 (79%) | 433/539 (80%) | 0.64 |
| **Use of antiplatelet drugs** | 1,589/1,813 (88%) | 1,105/1,274 (87%) | 484/539 (90%) | 0.070 |
| **Use of oral anticoagulants** | 205/1,807 (11%) | 156/1,270 (12%) | 49/537 (9.1%) | 0.053 |
| **Ipsilateral carotid stenosis<sup>3</sup>** |  |  |  | 0.021 |
| 0-50% | 10/1,779 (0.6%) | 7/1,245 (0.6%) | 3/534 (0.6%) |  |
| 50-70% | 143/1,779 (8.0%) | 114/1,245 (9.2%) | 29/534 (5.4%) |  |
| 70-99% | 1,626/1,779 (91%) | 1,124/1,245 (90%) | 502/534 (94%) |  |
| **Contralateral carotid stenosis<sup>3</sup>** |  |  |  | 0.41 |
| 0-50% | 949/1,662 (57%) | 661/1,171 (56%) | 288/491 (59%) |  |
| 50-100% | 713/1,662 (43%) | 510/1,171 (44%) | 203/491 (41%) |  |
| **Pre-procedural neurological symptoms** |  |  |  | 0.10 |
| *Asymptomatic* | 234/1,813 (13%) | 178/1,273 (14%) | 56/540 (10%) |  |
| *Ocular* | 334/1,813 (18%) | 223/1,273 (18%) | 111/540 (21%) |  |
| *TIA* | 754/1,813 (42%) | 523/1,273 (41%) | 231/540 (43%) |  |
| *Stroke* | 491/1,813 (27%) | 349/1,273 (27%) | 142/540 (26%) |  |
| **Percentage glycoprophorin C stain<sup>4</sup>** | 5.89 (2.69, 12.08) | 7.15 (3.37, 13.41) | 4.06 (1.98, 8.32) | <0.001 |

<sup>1</sup>Mean (SD); n/N (%); Median (IQR).<sup>2</sup>Wilcoxon rank sum test; Pearson's Chi-squared test; Fisher's exact test.<sup>3</sup>Degree of stenosis determined by US or CTA according to NASCET.<sup>4</sup>Calculated as (glycophorin C stain / plaque size)\*100.
BP = blood pressure; HDL= high density lipoprotein; LDL= low density lipoprotein; BMI= body mass index; eGFR = estimated Glomerular filtration rate; TIA = transient ischemic attack.

### Glycophorin C staining

Glycophorin C was used for staining and visualization of erythrocytes and erythrocyte membranes in culprit lesions of atherosclerotic plaque, revealing apparent differences between men and women (**Figure 2**). In addition, in certain samples with high glycophorin C staining, the distribution of erythrocytes was diffuse rather than concentrated at one specific location or hotspot in the plaque (**Figure 2**).

**Figure 1.**
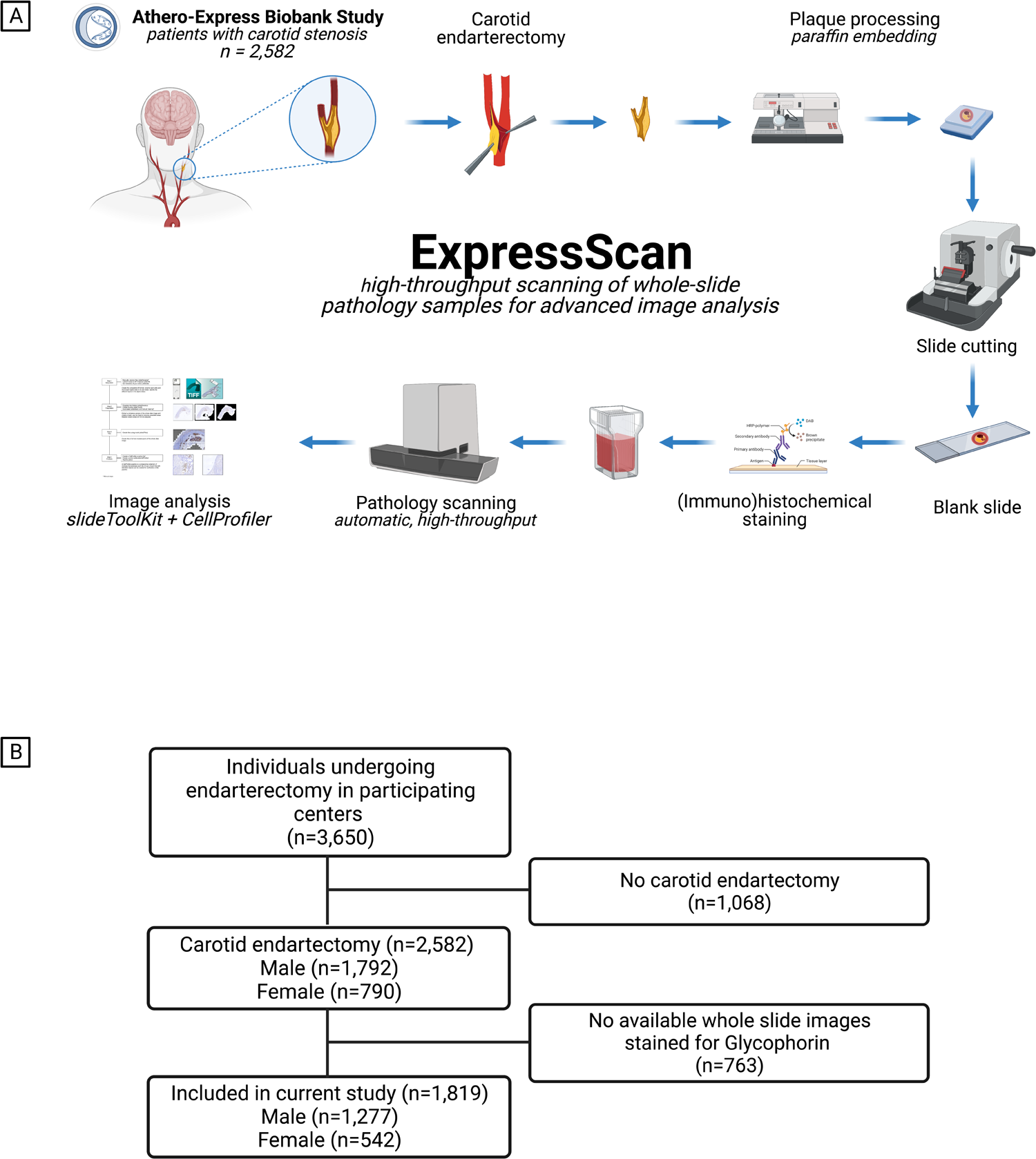
Study design and flowchart of study participants. **A.** Graphical illustration of the study design. Created with BioRender.com. **B.** Flowchart of number of individuals included in the analyses of the current study. A total of 1,068 patients were not included in the current study since they underwent iliofemoral endarterectomy. Additionally, 763 patients did not have available WSI’s with a glycophorin C staining.

**Figure 2.**
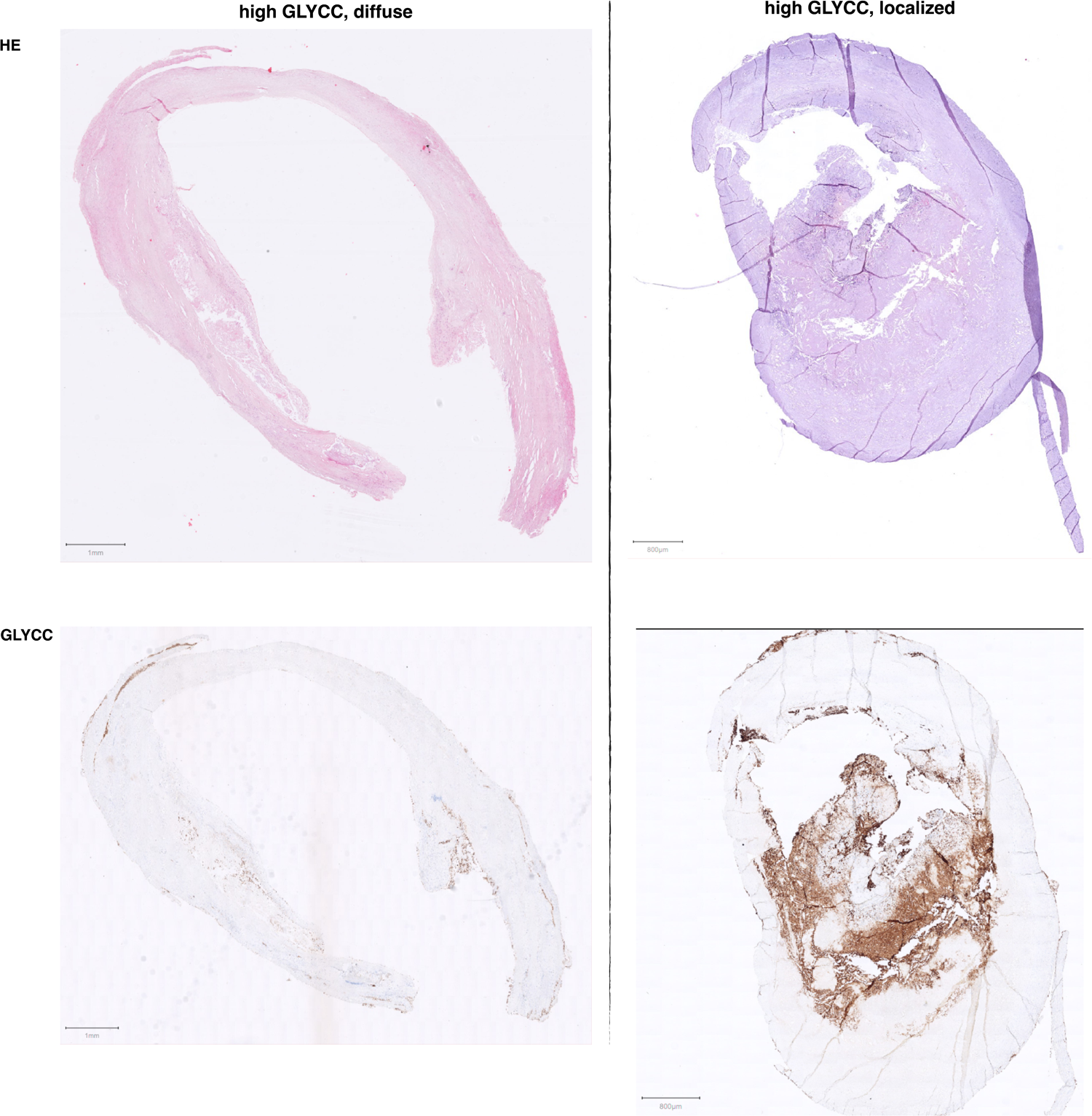
Example of slides with glycophorin C (GLYCC) and H&E staining. Displayed are two examples samples: left image is representative of a ‘diffuse’ glycophorin C staining in women, the right image shows a more ‘localized’ ‘core’-staining with some staining outside the ‘core’ in men. Be aware of the slight differences in scale and form between HE- and glycophorin C-stained samples from the same patients, these are sequential cuts.

### Glycophorin C and intraplaque hemorrhage

In general, plaques with greater percentage total plaque area covered with glycophorin C, were plaques showing intraplaque hemorrhage as scored semi-quantitatively (3.70 (IQR: 1.74, 7.27), 6.99 (IQR 3.12, 13.26), p<0.001, **Figure 3, Methods**). However, some outliers were observed that contained a large area stained for glycophorin C but were previously scored as plaques without IPH using semi-quantitative staining method. We also observed this in the histological slides stained for glycophorin (**Figure 4**).

**Figure 3.**
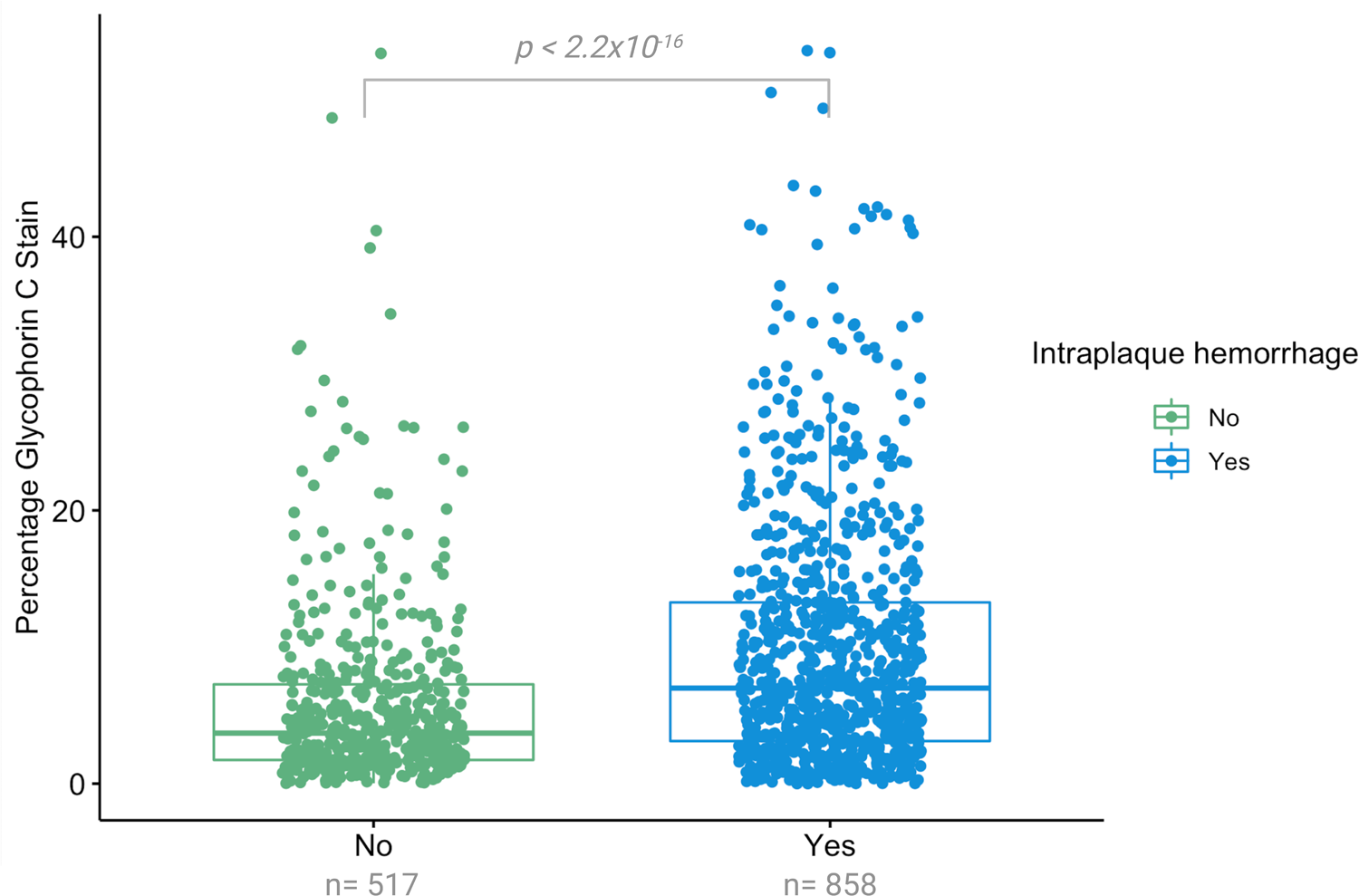
Univariable association between glycophorin C expressed as percentage glycophorin of total plaque surface with presence of intraplaque hemorrhage (IPH) P-values are derived from Kruskal Wallis test. Shown are the median values (central line), the upper and lower quartiles (box limits), and the 1.5x interquartile range (whiskers). The percentage of glycophorin C (inverse rank transformed) was significantly higher in samples with intraplaque hemorrhage in comparison to samples without IPH (p<0.001).

**Figure 4.**
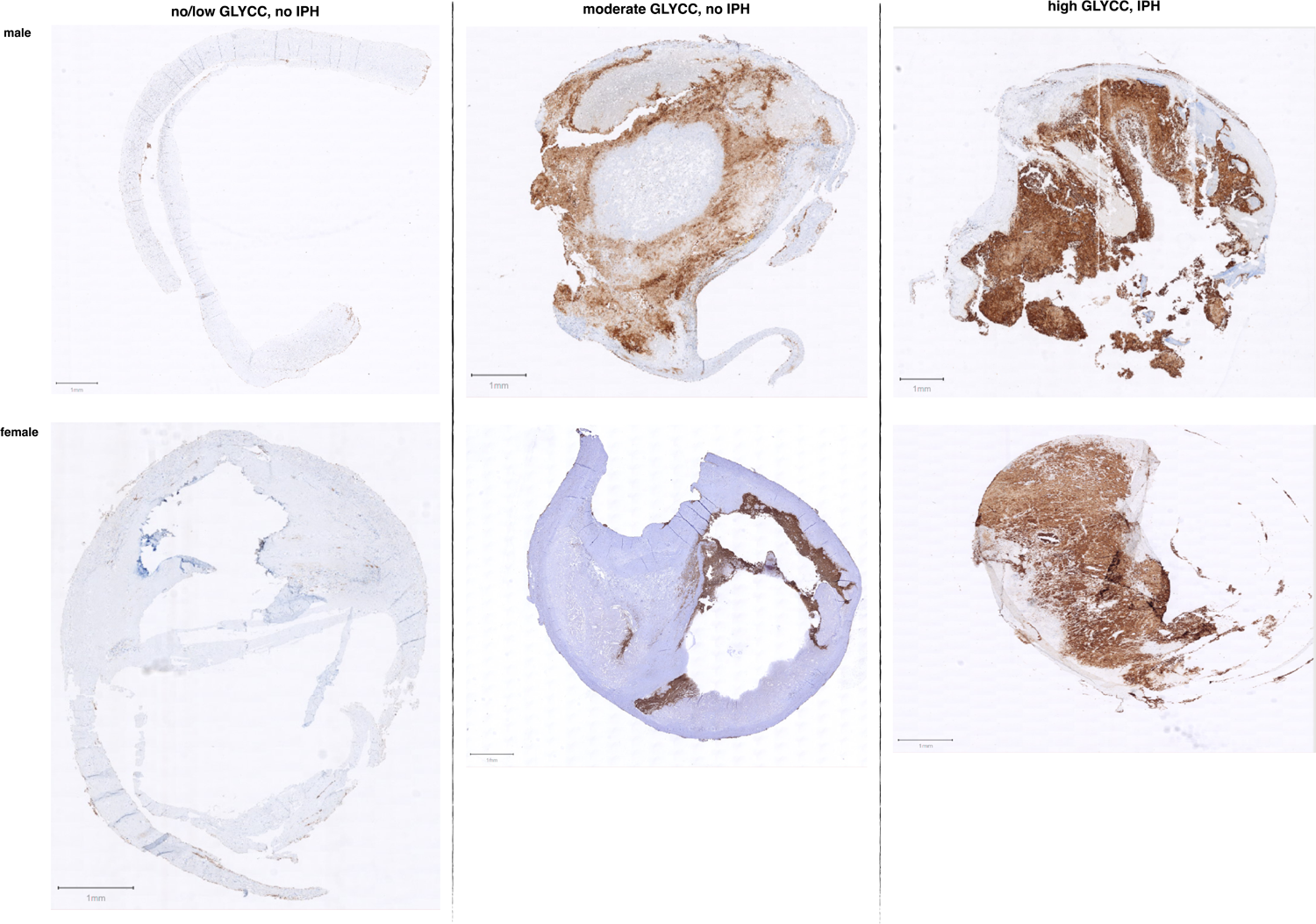
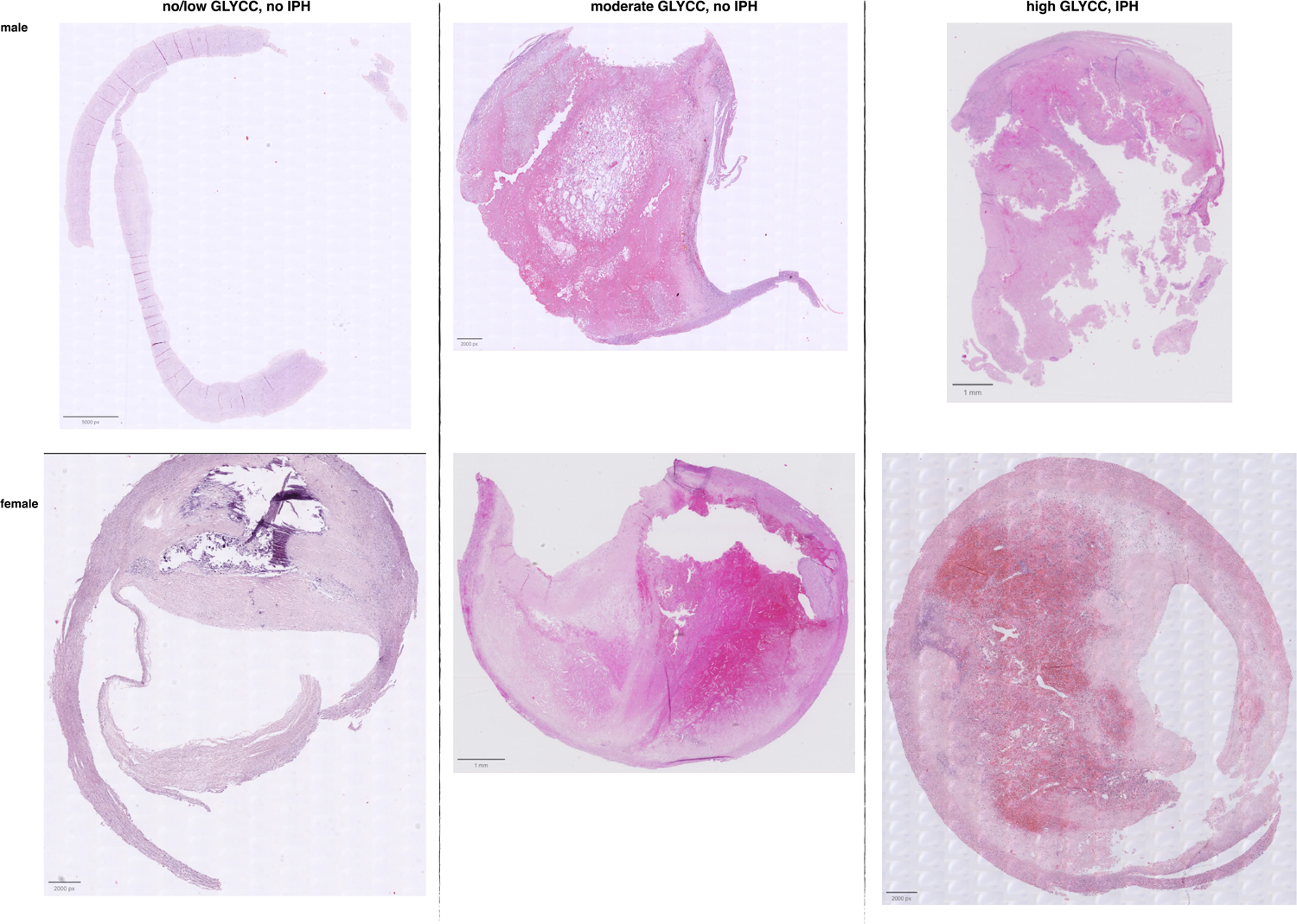
Example of slides with different levels of glycophorin C (GLYCC) staining compared to the presence of intraplaque hemorrhage (IPH) Displayed are example 6 individual patient samples for low, moderate and high glycophorin C (upper panel) staining in samples from patients in which IPH was scored as present or not present (displayed a HE stain, lower panel). Be aware of the slight differences between HE- and glycophorin C-stained samples from the same patients, these are sequential cuts.

Glycophorin C, defined as the total plaque area that is positive for glycophorin C, adjusted for total plaque size was associated with the presence of IPH in atherosclerotic plaques (Model 1: OR 1.83; 95% CI 1.59, 2.12; p=<0.001, **Figure 5A**). After adjustments for cardiovascular confounders the association remained significant (Model 2: OR 1.90; 95% CI 1.63, 2.21; p=<0.001). Separate sex-stratified analyses adjusted for confounders, showed significant association of glycophorin C with IPH in both men (Model 2: OR 1.85; 95% CI 1.53, 2.26; p=<0.001) and in women (Model 2: OR 1.43; 95%CI 1.12, 1.84; p=0.004, **Figure 5A**).

**Figure 5.**
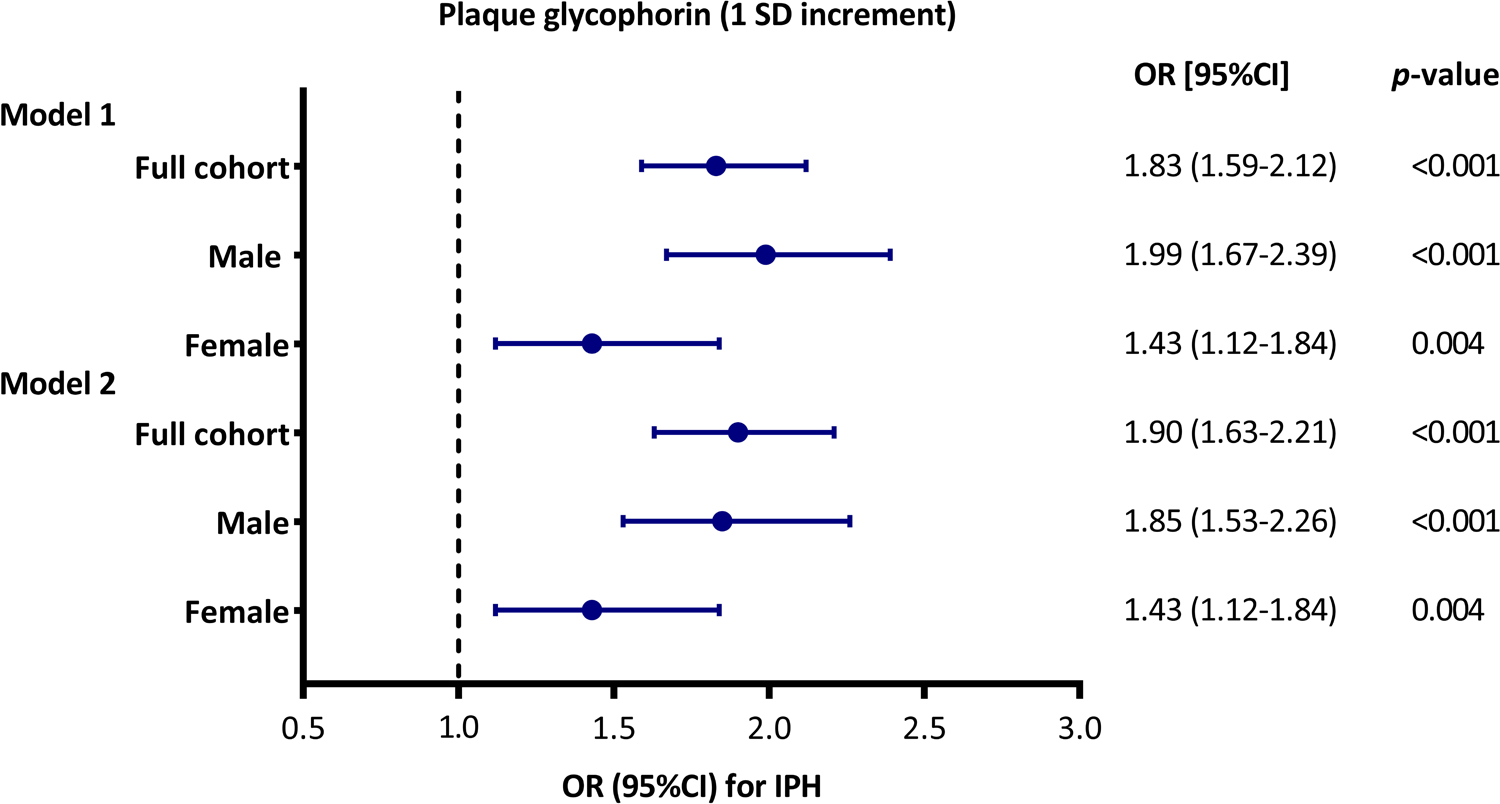
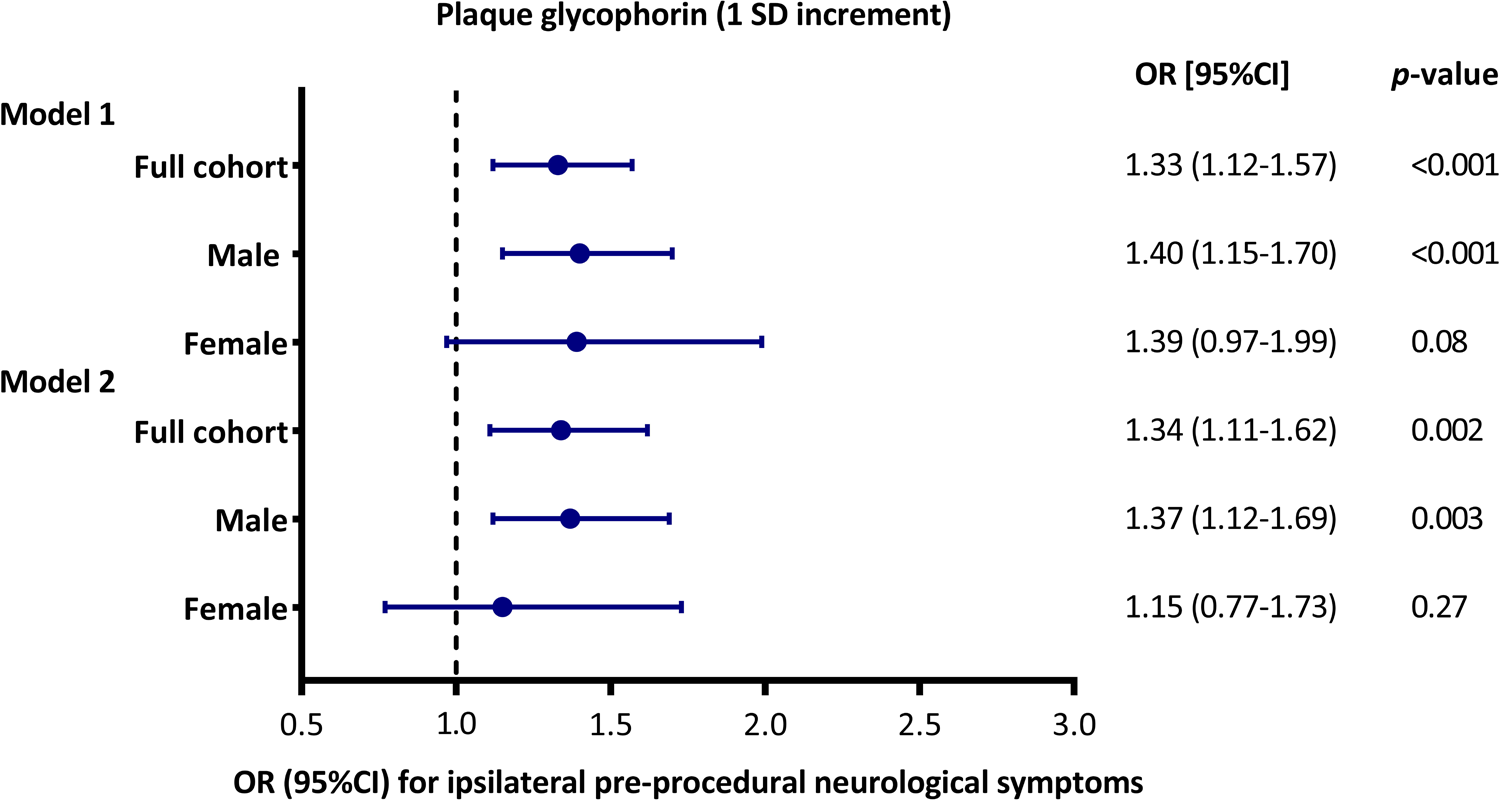
Multivariable associations between plaque glycophorin C (1 SD increment) with IPH and pre-procedural neurological symptoms. Associations adjusted for plaque size (Model 1) and adjusted for plaque size and confounders (Model 2) are shown. Shown are odd ratios (OR) or hazard ratios (HR) and error bars corresponding to their 95% confidence intervals (CI). The confounders included in the model 2 associations can be found in **Supplemental Table 2. A.** Multivariable associations of glycophorin C with IPH, as derived from logistic regression analyses. **B.** Multivariable associations of glycophorin C with pre-procedural symptoms (asymptomatic versus symptomatic), as derived from logistic regression analyses.

### Glycophorin C and pre-procedural neurological symptoms

Overall, increased staining of glycophorin C, corrected for plaque size, was associated with neurological symptoms before CEA (Model 1: OR 1.33; 95%CI 1.12, 1.57; p<0.001), the association remained significant after correction for multiple cardiovascular confounders (Model 2: OR 1.34; 95%CI 1.11, 1.62; p=0.002) (**Figure 5B**). In the separate sex-stratified analysis, plaque size corrected glycophorin C was found in men but not in women (Model 2: OR 1.37; 95%CI 1.12, 1.69; p=0.003 and Model 2: OR 1.15; 95%CI 0.77, 1.73; p=0.27, respectively) (**Figure 5B**).

### Glycophorin C and overall plaque phenotype

The total plaque surface covered by glycophorin C expressed as a percentage of the total plaque size was associated with the overall plaque phenotype. Overall, a gradual increase in glycophorin C staining was seen in fibroatheromatous and atheromatous plaques in comparison to fibrous plaques (**Figure 6, Supplemental Table 3)**. The percentage of glycophorin C was significantly higher in fibroatheromatous and atheromatous plaques in comparison to fibrous plaques (*p*=8.8×10^-12^ and *p*<2.10×10^-16^, respectively, **Figure 6**). Additionally, the percentage of glycophorin C was significantly higher in atheromatous plaques in comparison to fibroatheromatous plaques (*p*=3.4×10^-8^, **Figure 6**).

**Figure 6.**
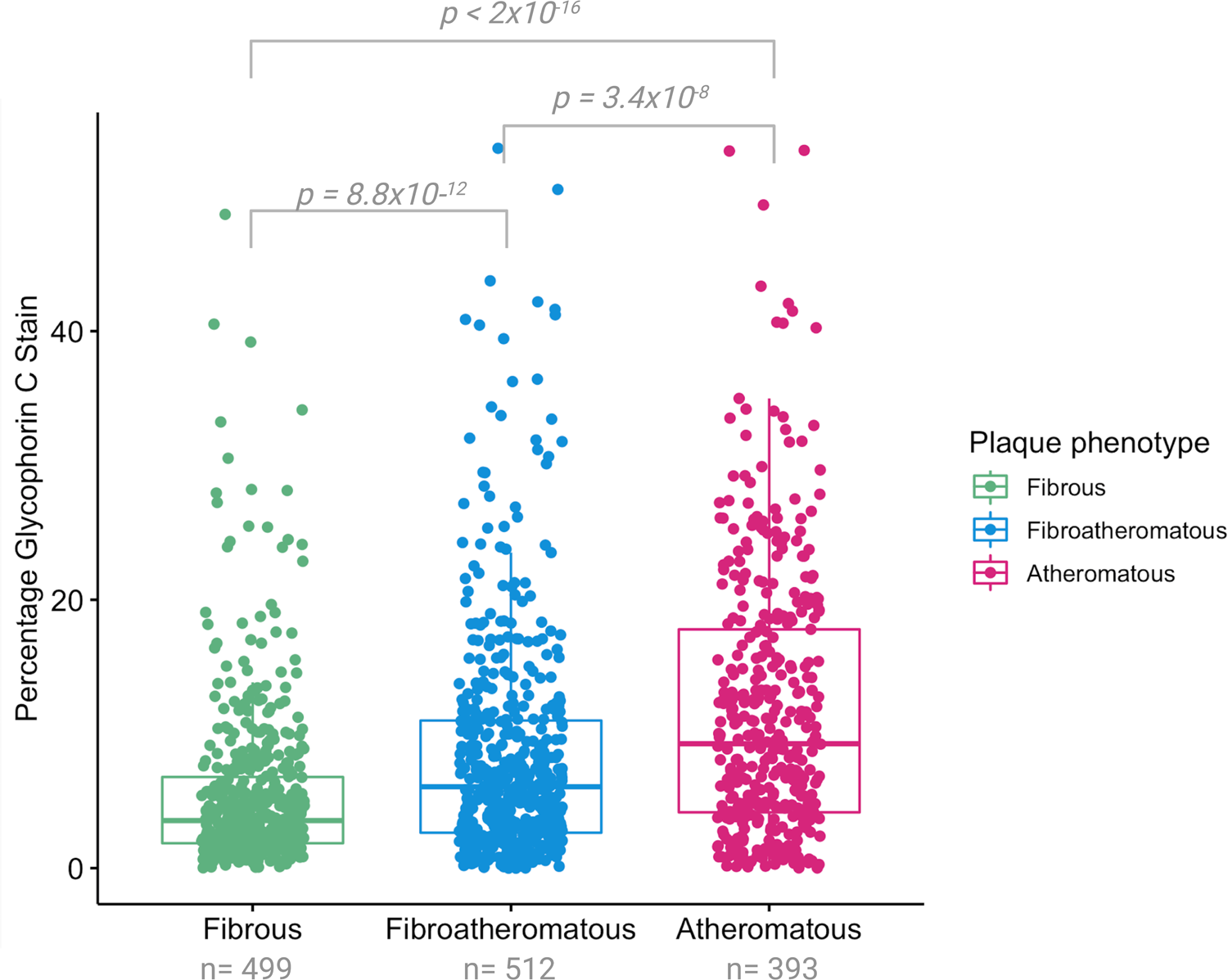
Univariable association between glycophorin C expressed as percentage glycophorin of total plaque surface with overall plaque phenotype. P-values are derived from pairwise comparisons using Wilcoxon rank sum test. Shown are the median values (central line), the upper and lower quartiles (box limits), and the 1.5x interquartile range (whiskers). The percentage of glycophorin C (inverse rank transformed) was significantly higher in fibroatheromatous and atheromatous plaques in comparison to fibrous plaques. Additionally, the percentage of glycophorin C (inverse rank transformed) was significantly higher in Atheromatous plaques in comparison to fibroatheromatous plaques.

### Associations with histological plaque characteristics

Besides its strong correlation with IPH, glycophorin C adjusted for plaque size was also associated with a large lipid core (OR 1.85; 95%CI 1.60, 2.15; p<0.001) and a higher macrophage burden (OR 1.87; 95%CI 1.63, 2.14; p<0.001, **Figure 7**). In addition, but less calcifications (OR 0.81; 95%CI 0.71, 0.91; p<0.001), and a lower collagen content (OR 0.70; 95%CI 0.60, 0.82; p<0.001) and lower SMC content (OR 0.60; 95%CI 0.52, 0.68; p<0.001) (**Figure 7**). The separate sex stratified analysis identified apparent sex differences in the associations of glycophorin C with large lipid core, calcifications, collagen content, macrophage burden and IPH (**Figure 7**). In women glycophorin C was not associated with calcifications and collagen content (OR 1.07; 95%CI 0.85,1.35; *p*=0.57 and OR 0.83; 95%CI 0.62,1.12; *p*=0.21, respectively) while in men the associations were similar to the full cohort (**Figure 7**). In addition, the effect sizes of the association between glycophorin C and macrophage burden and IPH were lower in women compared to men (OR 1.45; 95% 1.14,1.86 *p*=0.003 vs. OR 2.01; 95% 1.70, 2.39 *p*<0.001 and OR 1.43; 95% 1.12,1.84 *p*=0.005 vs. OR 1.99; 95% 1.67, 2.39 *p*<0.001, respectively) (**Figure 7**).

**Figure 7.**
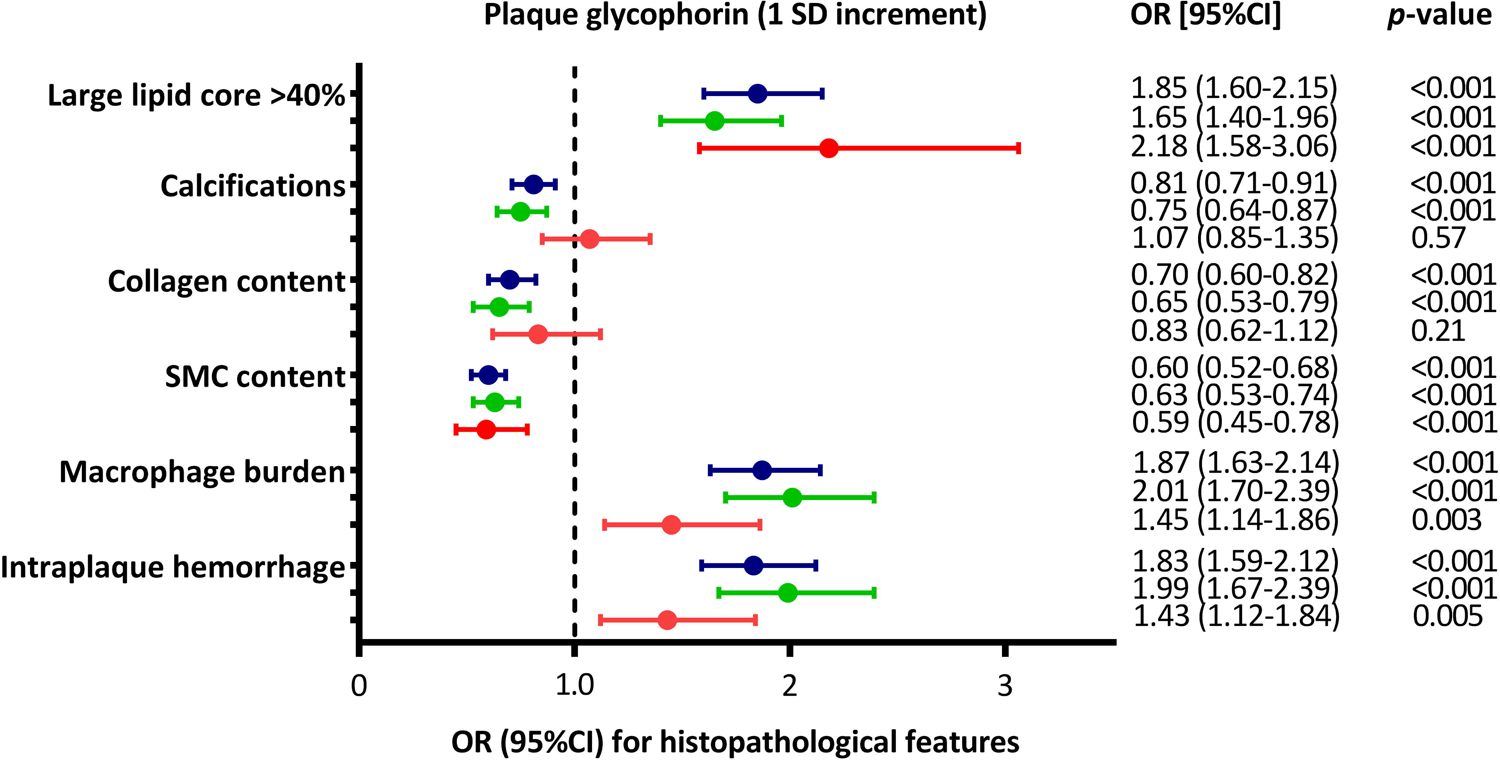
Association of plaque glycophorin C with (1 SD increment) with the individual (semi)-quantitative histopathological features (binary traits) Shown are odds ratios (OR) and error bars correspond to their 95% CI, as derived from logistic regression analyses. Blue error bars are the full cohort, and the green and red error bars are male and female subset, respectively.

### Glycophorin and major adverse cardiovascular events

Glycophorin C was not associated with MACE in Cox regression analysis adjusted for plaque size (Model 1: HR 1.15; 95%CI 0.98, 1.35; p=0.09) nor in a model adjusted for plaque size and confounding baseline variables (Model 2: HR 1.14; 95%CI 0.95, 1.35; p=0.16, **Supplemental Figure 1**). Sex-stratified analyses also showed no association between glycophorin C and future MACE (**Supplemental Figure 1**). Interestingly, semi-quantitatively scored intraplaque hemorrhage was associated with MACE (Model 1, **Supplemental Figure 2**). After adjusting for confounders, the association between IPH and MACE remained significant (**Supplemental Figure 2**), and sex-stratified analyses showed a significant association in men but not in women (**Supplemental Figure 2**).

## Discussion

We demonstrated that visualized and quantified glycophorin C, a marker of the accumulation of erythrocytes within atherosclerotic plaque, was associated with the IPH, a potent marker for future MACE in patients that underwent CEA^15,16^. Furthermore, after adjusting for cardiovascular confounders glycophorin C was associated with pre-procedural neurological symptoms. Additionally, glycophorin C was associated with classical features of vulnerable plaques^17^, such as a larger lipid core, a higher macrophage burden, a lower collagen content and a lower SMC content. Notably, the amount of glycophorin C staining gradually increased in more advanced plaque phenotypes, which further strengthens the notion that quantifying erythrocyte accumulation by glycophorin C can be used a marker for plaque vulnerability.

Several mechanisms have been described to explain how erythrocytes accumulate in plaque, including via plaque fissures^18^, incorporation of a luminal erythrocyte-rich thrombus into the plaque^19^ or, most likely, via leaky newly-formed microvessels^20^. Microvessels are lined with discontinuous abnormal endothelial cells that allow the extravasation of leukocytes and erythrocytes^21^. The pro-inflammatory and pro-oxidative environment makes the erythrocytes vulnerable to hemolysis or phagocytosis by intraplaque macrophages and SMC^22^, leading to the release of erythrocyte components such as cholesterol, iron, heme and hemoglobin molecules. Due to the high cholesterol content of erythrocyte membranes, of which lipids account for 40% by weight^5^, erythrocyte membranes can deliver a significant amount of lipids to the necrotic core. Iron, heme and hemoglobin can trigger processes that contribute to plaque instability, including endothelial activation and dysfunction, SMC proliferation and migration, foam cell formation, platelet aggregation and macrophage activation^23–25^. Together, the accumulation of erythrocyte membranes and subsequent processes within an atherosclerotic plaque represent a powerful atherogenic stimulus. This is confirmed in an animal experiment in which autologous erythrocytes were injected directly into stable atherosclerotic lesions in the aorta of rabbits^4^. Erythrocyte injection triggered the enlargement of necrotic core and formation of free cholesterol crystals along with excessive macrophage infiltration^4^. Our results, derived in a human population, support this observation as glycophorin C was associated with a larger lipid core and more intraplaque macrophages. Lysed erythrocytes, particularly their membranes, but not intact erythrocytes, contribute to atherosclerosis progression by enhancing vascular calcification^26^. However, in our current study, we observed an inverse relationship between glycophorin C and calcifications within the plaque. This discrepancy can be attributed to both the study protocol preceding histological staining and our quantification method for glycophorin C. The standardized study protocol involves the decalcification of plaques before histological staining. As a result, the visual assessment of the calcification phenotypes relis on Hematoxylin and Eosin (H&E) staining, which allows the evalualtion of tissue-free areas in the plaques. However, H&E staining is not specific to erythrocytes and cannot differentiate erythrocyte membranes. It is commonly employed to examine general tissue structure, determine the extent of hemorrhage, and classify IPH using a semi-quantitative method based on intact erythrocytes or hemorrhagic debris. The latter consists of a mixture of intact and degenerated erythrocytes, fibrin, amorphous material, and myofibroblasts located in the core of the plaque^11,27^. To differentiate between old IPH with erythrocyte membrane remnants and new IPH with intact erythrocytes, additional staining methods would be required. For instance, Perl’s staining can visualize free iron in the tissue, but it lacks specificity as it also detects iron deposits in other structures such as macrophages and foam cells^1^. Combining stains like H&E, glycophorin C, and Perl’s in a histological stratification approach at a larger scale could provide valuable insights in future studies. Nevertheless, our current findings offer compelling evidence that quantifying erythrocyte accumulation through glycophorin C staining, may serve as a marker for the evolution of atherosclerosis towards a plaque with a ‘vulnerable’ phenotype. While further investigations incorporating comprehensive histological stratification methods are warranted, our findings provide sufficient support for the potential significance of glycophorin C staining as an indicator of atherosclerotic progression.

In the current study, we found an association between sex with quantified glycophorin C. Similarly, we found that glycophorin C was only associated with pre-procedural neurological symptoms in men but not in women. We hypothesize that the differences in the association between glycophorin C and pre-procedural neurological symptoms in men and women might be explained by the differences in plaque composition. The underlying mechanism that explains sex-related differences in plaque composition is yet to be unraveled, however evidence that the plaque composition differs between men and women is accumulating^28^. Iliac-femoral plaques from men had a higher prevalence of lipid cores and plaque hemorrhage compared to women^29^. A large coronary computed tomography angiography (CTA) study revealed that plaques from men had a higher prevalence of high-risk plaque features on CTA and were more heavily calcified^30^, the latter was also found in autopsy study^31^. In the sex-stratified analysis of the associations of glycophorin with semi-quantitative scored plaque characteristics, we also found marked differences between men and women. Glycophorin C was not associated with calcifications and collagen in women. This might be due to lower levels of glycophorin C in women as demonstrated in the current paper, or the possibility that plaques of women generally contain more calcifications or collagen^28^. The differences in plaque composition between men and women may account for variations in the relationship between glycophorin C and pre-procedural neurological symptoms. The association of glycophorin C with high-risk plaque features in men suggests a potential explaination for localized prothrombotic properties. The presence of pre-procedural neurological symptoms and the occurrence of MACE can be attributed to heightened procoagulant activity, often triggered by plaque rupture^32^. The accumulation of erythrocytes within atherosclerotic plaque plays a significant role in this process, acting as a critical trigger for procoagulant mechanisms.

Early stage and advanced atherosclerotic lesions express Tissue factor (TF), a major initiator of the blood coagulation cascade, contributing to the thrombotic nature of human atherosclerotic lesions^33^. Notably, TF activity is significantly higher in lesions from patients with unstable cardiovascular disease compared to those with stable forms of cardiovascular disease^34^. Futher investigation is required to determine whether glycophorin C quantification, indicative of erythrocyte accumulation, correlates with increased TF activity and heightened plaque thrombogenicity.

Although glycophorin C has been strongly linked to IPH, our study did not find glycophorin C to be an independent predictor of future cardiovascular events. However, in a subsequent post-hoc analysis utilizing more samples, we able to replicate our previous findings, demonstrating that IPH is indeed a predictor of future events^14^. Additional, we previously observed a significant association between sex and IPH^14^, with a higher prevalence of IPH among men compared to women. This finding aligns with our observation that samples obtained from men exhibit significantly higher levels of glycophorin C. The location of IPH within a plaque is considered while semi-quantitatively scoring IPH, but the location of staining is not considered in the current glycophorin C quantification method. We hypothesize that plaques with diffuse staining of glycophorin C may not be as vulnerable as those with more concentrated staining near neo-vessels or at more vulnerable locations within the plaque, which could be related to sex (**Figure 2**). Additionally, we found discrepancies between automated and manual assessments of glycophorin C and IPH, suggesting that manual assessment may be prone to observer variability and human error.

There are a few limitations in the present study. First as the Athero-Express Biobank represents a retrospective study design, we can only speculate about the causality of our findings. Second, as discussed in our previous work, the term “intraplaque hemorrhage” might not be appropiate, as endarterectomy specimens are frequently fragmented, making it difficult to determine the exact source of plaque hemorrhage. They might be due to plaque bleeding at the luminal site due to plaque disruption or as a result of defective leaky micro vessels^15,35^. However, the hemorrhages revealed signs of organization during assessment, this did indicate that the plaque hemorrhage are not artifacts of surgical dissection^15^. Third, as part of the study-protocol plaques are decalcified, thus ‘calcification’ is manually assessed using H&E and based on remnant tissue-free areas in the plaques. The ‘calcification’ phenotype serves as a rough approximation and has limited utility in accurately evaluating the extent. Additionally, the `slideToolKit` quantitatively scores the positively stained plaque areas regardless of their location within the plaque, these areas can be clustered in hotspots or have a diffuse distribution within the plaque. While this may also be an advantage, knowing the location and intensity of stained areas for glycophorin C might provide additional value in relation to other histological plaque features and clinical symptomatology. We hypothesize that diffuse staining is related to old IPH, healed ruptures or thrombi and thus to previous symptoms, and that hotspots may indicate areas that are rich in neovascularisation, which may be related to future events. This stratification could be highly relevant, however, within the current glycophorin C `Cellprofiler`-pipeline in `slideToolKit` (Supplementary Figure 3), this is not yet possible^9^. Next, the quantitative assessment of the plaque characteristics was based on the analysis of a single WSI, rather than a serial assessment of various WSIs throughout the plaque. Based on previous research, we deem it unlikely that this strongly influences our results, as there was good overall agreement between three-to five-millimeter sections in histological assessment studies using the semi-quantitative classification method^11,12^. Indeed, depending on the methodology used and the specific feature of the atherosclerotic plaque under evaluation, variations may occur. Even so, when investigating the plaque composition between patient groups, a large cohort with less extensive sampling per plaque would be sufficient^12^. This minimizes chance findings and allows for statistical analysis to correct for confounding factors. Large studies such as this, would be therefore unlikely to miss differences between patient groups due to current variation levels^12^. In fact, the inclusion of precise quantification using `CellProfiler` within our `slideToolKit` workflow has enabled the identification of a plaque instability feature that genuinely reflects plaque development. Notably, this approach offers the distinct advantage of avoiding subjective scaling (such as high, medium, or low) and instead provides an exact, accurate, and robust quantification. Thus, by adopting this methodology, we have enhanced the reliability and objectivity of our assessment, ensuring a more rigous and compehrensive evaluation of plaque instability. In future investigations, it would be valuable to explore potential intersegment variations in glycophorin C levels, specifically by comparing the shoulder region with the culprit lesion. This targeted analysis would provide insights into any disparities in glycophorin C levels as marker of (remnant) erythrocytes within different regions of the same plaque, potentially shedding light on distinct pathophysiological mechanisms and their contribution to plaque instability. Such an investigation would further enhance our understanding of the role of erythrocytes and intraplaque bleeding and its localization within atherosclerotic plaques, offering valuable insights for clinical risk stratification and targeted therapeutic interventions. Another point that needs to be addressed is that the concept around the ‘vulnerable’ plaque has evolved, and the classification of atherosclerotic plaques is more complex than previously thought^36^. The diversity and complexity of lesion types go beyond the perceived overall plaque phenotypes, thus the presence of one specific plaque feature probably does not determine outcome. Furthermore, although we described one of the largest groups of women operated by CEA, confounding in the multivariable analysis may have been impeded by power, i.e., sample size. These sex-stratified results need to be interpreted in the light of the confidence intervals, to avoid misleading conclusions^37^. In addition, for the longitudinal analyses we have used MACE as a composite outcome, and we were not able to perform analyses on separate outcomes such as (non)fatal stroke or non(fatal) myocardial infarction due to a limited number of events.

A noteworthy strength of our study lies in the demonstration of the informative value of glycophorin C staining of erythrocytes and erythrocyte remnants in assessing plaque stability. Traditionally, IPH has been evaluated using a binary scoring system, indicating its presence or absence. In contrast, the quantification of glycophorin C represents a continuous measure of erythrocytes accumulation within atherosclerotic plaque, offering the opportunity to interpret the severity of IPH. Consequently, by employing quantified glycophorin C, we can obtain more comprehensive insights into the extent and severity of IPH within the plaque, thereby aiding in the characterization of high-risk plaques prone to instability. This enhanced level of information distinguishes quantified glycophorin C as a superior method for measuring IPH and identifying plaques with potential for instability. At present, glycophorin C, as a proxy for erythrocyte accumulation, is promising as a tool for improving risk stratification, but further research is needed to determine its potential clinical utility. Moreover, this is the first study in which we apply the fully automated `slideToolKit` workflow at scale for a specific staining. As such, this study confirms the effectiveness of our `slideToolKit` workflow, allowing further reliable determination of plaque composition and the exploration of possible new histological (bio)markers. Our `CellProfiler`-based workflow and results can used, in large scale translational ‘-omics’ studies and for the validation of non-invasive imaging techniques, both of which heavily rely on accurate, robust and reproducible histological scoring methods as the gold standard comparison^38^.

In summary, the accumulation of erythrocytes visualized and quantified by glycophorin C was independently associated with IPH and other features of a more vulnerable plaque in both men and women. In addition, in men undergoing CEA glycophorin C was independently associated with pre-procedural neurological symptoms. These findings support the notion that staining erythrocytes and erythrocyte remnants with glycophorin C provides valuable information about plaque stability, especially for men.

## Materials and methods

### Study population

We used culprit lesions of human atherosclerotic plaques collected in, and data from, the Athero-Express Biobank (**Figure 1B**), an ongoing prospective study of patients undergoing carotid endarterectomy (CEA) for atherosclerotic carotid stenosis. A detailed description of the study design has been published previously^39^. In short, patients were recruited from the St. Antonius Hospital Nieuwegein, Nieuwegein, the Netherlands and University Medical Center Utrecht in Utrecht, the Netherlands from 2002 onwards. During the longevity of the Athero-Express biobank the indications for CEA were reviewed by a multidisciplinary vascular team according to the latest recommendations of the ESVS guideline^40^. Individuals who agreed to participate completed standardized baseline questionnaires regarding cardiovascular risk factors, medical history and medication use prior to surgery, that were verified against medical records. Pre-operative blood samples were collected, processed and stored for future use. For this study, all participants that underwent CEA, with available WSI’s stained for glycophorin C, were included (**Figure 1**). Plaque samples were freshly obtained during surgery and analyzed as described below. The current study was conducted in accordance with the Declaration of Helsinki and was approved by the ethical boards of both hospitals. All patients provided written informed consent.

### Atherosclerotic plaque processing

CEA derived plaque samples were immediately transferred and processed (**Figure 1A**). The plaque was divided in parallel segments of 5-mm thickness perpendicular to the arterial axis and the segment with the greatest plaque burden, the culprit lesion, was subjected to histopathological examination, as previously described^12,15^. In short, the culprit lesion was washed to remove presumed artifacts of the surgery such as red blood cells surrounding the dissection planes and fixed the culprit lesion in 4% formaldehyde, subsequently decalcified, embedded in paraffin and cut into 5 μm sections that were routinely stained for different characteristics, which was scored manually by a single experienced observer as no/minor or moderate/heavy staining according to predefined criteria using a microscope with 40x magnification^15^. One section per staining per patient was scored. The Hematoxylin-Eosin (H&E) was used to obtain a general overview to assess any calcification(s) and the lipid core, picrosirius red and elastin von Gieson stainings were used to score collagen, the alpha-actin staining was used for smooth muscle cells (SMCs) and the CD68 staining for macrophages. The presence of intraplaque hemorrhage (IPH) was defined as hemorrhage within the plaque tissue and was rated as being absent or present using H&E and Fibrin (Mallory’s phosphotungstic acid-hematoxylin) staining. The overall plaque phenotype was scored as either, fibrous, fibroatheromatous and atheromatous based on the overall appearance of the plaque by the same experienced observer, which is mainly based on the percentage of plaque that is occupied by lipid^39^. Intra-observer and interobserver variability have been examined previously and showed moderate to good reproducibility (κ 0.6-0.9) for the semi-quantitative scoring method^12^.

### Glycophorin staining

We used glycophorin C because this antibody has been routinely used in the automatic staining machine for immunohistochemistry of the diagnostic pathology laboratory of the University Medical Center Utrecht in hematopathology diagnostics. In this way we could guarantee that the staining was reproducible and has the same staining conditions in all slides of the study population. Although glycophorin A is more abundant with 5-9×10^5^ copies, than the 0.5-1.0×10^5^ copies of glycophorin C per cell, both give the same staining pattern of (remnants of) erythrocyte membranes due to the high prevalence of copies^41^. To visualize glycophorin C, sections were stained with a monoclonal mouse anti-Glycophorin C antibody (clone Ret40f, DAKO, dilution 1:800) using the Ventana BenchMark Ultra system (Roche Diagnostics) with CC1 (EDTA) pretreatment during 24 minutes. The plaque characteristic glycophorin C was defined as the total area positively stained, adjusted for the total plaque size in μm^2^.

### ExpressScan: histopathological analysis of atherosclerotic plaque characteristics

We used the `slideToolKit`^9^ to quantify the amount of glycophorin C staining on culprit lesions from carotid atherosclerotic plaque samples. The `slideToolKit` workflow has been published previously and involves a four-step process using a collection of open-source scripts to support the analyses of WSIs^9,13^. WSIs are slides that are completely scanned at high magnification and stored as digital images, also known as WSI or digital slides. In short, in step one (*acquisition*), the WSIs were obtained and stored. To this end, we set up the ExpressScan to scan all stained sections using a Roche Ventana iScan HT slide scanner or a Hamamatsu C12000-22 Digital slide scanner and saved brightfield microscopy WSIs as z-stacked TIF (Ventana) and NDPI (Hamamatsu), sized ∼300Mb to ∼1.1Gb each. In the second step (*preparation*), the files of a give stain were organized into stain-folders and all individual images *sub*folders within that stain-folder. To increase downstream analytical efficiency, the non-tissue parts of the images were masked (blackened) to enable exclusion during the next two steps. In the third step, the WSIs were divided into tiles (tiling), which created tiled images of manageable size for later analysis. In the fourth and final step (analysis), the tiled WSIs were analyzed using custom pre-trained pipelines (protocols) using `CellProfiler` v4.1.3N^9^; results were stored and combined in a comma-separated dataset. For glycophorin C a `CellProfiler`-pipeline was created to assess and quantify the total area positively stained, adjusted for the total plaque size in μm^2^. Note that the histological cutting procedure may result in multiple section on one slide; the slide scanner is indifferent to this and thus WSI may contain multiple section. As we adjust the total area positively stained for the total area measured across sections – whether that is one or multiple is inconsequentional. This pipeline is available in the `slideToolKit` repository in the folder `pipeLines`. See **Supplemental Figure 3** for the glycophorin C `CellProfiler`-pipeline workflow using a single tile as an example. The main strengths of the `slideToolKit` method are that it is very scalable, fully automated, precise and scores excellent regarding reproducibility with intraclass correlation coefficients (ICCs) of 0.92-0.97^13^.

### Ipsilateral pre-procedural neurological symptoms

Patients were classified with either an ipsilateral asymptomatic or symptomatic carotid stenosis prior to surgery which was based on the review of their medical record. Patients without any ipsilateral symptoms in the last six months were classified as asymptomatic. Cerebrovascular events included ischemic stroke, a transient ischemic attack (TIA), amaurosis fugax, ocular ischemic syndrome or a central retinal artery occlusion.

### Follow-up analysis for major adverse cardiovascular events (MACE)

MACE was defined as the three-year postoperative risk of major adverse cardiovascular events including fatal or nonfatal ischemic or hemorrhagic stroke, fatal or nonfatal myocardial infarction (MI) and any vascular death, defined as death of presumed vascular origin. Clinical endpoints were assessed at one, two and three years after surgery by means of patient questionnaires and validated by review of medical records; if necessary, general practitioners or other treating physicians were contacted for follow-up data.

### Statistical analysis

Continuous variables were summarized as mean and standard deviation (SD) or as median and interquartile range (IQR). Categorical variables were expressed as frequencies and percentages. Glycophorin C and the total plaque size were inverse-rank transformed to standardize the data. Continuous data was assessed for skewness and natural log transformed in analysis when needed. Logistic regression analyses were performed to test the association between glycophorin and IPH, pre-procedural neurological symptoms and binary plaque characteristics. For the association between glycophorin and secondary MACE Cox proportional hazard models were performed. We have sex stratified the main results.

For the logistic regression models and Cox proportional hazard models (`Survival` package in R) we made two different models according to a specific structure, Model 1 adjusted for plaque size, Model 2 adjusted for plaque size and (potential) confounders. The confounders were identified by exploring the association of baseline characteristic, such as risk factor and medication use, with glycophorin adjusted for plaque size, and with the different dependent outcome variables (e.g., IPH and MACE). When the baseline characteristic was associated with the quantified plaque characteristic and with the outcome variable with a p-value of <0.1, the specific baseline characteristic was included as a potential confounder in the final logistic regression or Cox proportional hazards model, respectively. Across the selected potential confounders, the proportion of missing values was analyzed (**Supplemental Table 3**). Missing values of potential confounders were imputed using multivariate imputation by chained equations^42^(`mice` package in R). When correlated predictors were included in the final model, the best model was selected based on the Akaike Information Criterion (AIC).

Statistical significance threshold was set at a two-sided *p*-value <0.05 across all analyses.

Analyses were performed using R (v4.0.5; The R Foundation for Statistical Computing).

## Data and code availability

The data as used here are available through DataverseNL. DOI: https://doi.org/10.34894/08TUBV All documented code can be found here: https://github.com/CirculatoryHealth/ExpressScan_Glycophorin; the codes and scripts for slideToolKit are available here: https://github.com/swvanderlaan/slideToolKit.

## Author contributions

Study design: JM, DK, GB, SH, SWvdL

Data collection: JM, ND, MV, SWvdL, GP

Sample measurements: ND, YS, CM, SWvdL

Data analysis and interpretation: JM, TS, MV, DK, MM, HR, GP, YS, CM, AF, SH, SWvdL

Drafting the article and figures: JM, MV, DK, GB, SH, SWvdL

Critical revisions and final approval: JM, TS, MV, ND, YS, CM, AF, GP, MM, HR, DK, GB, SH, SWvdL

## Data Availability

The data as used here are available through DataverseNL. DOI: https://doi.org/10.34894/08TUBV

https://doi.org/10.34894/08TUBV

## Acknowledgements

We would also like to thank all the (former) employees involved in the Athero-Express Biobank Study of the Departments of Surgery of the St. Antonius Hospital Nieuwegein and University Medical Center Utrecht for their continuing work. Lastly, we would like to thank all participants of the Athero-Express Biobank Study; without you these kinds of studies would not be possible.

