## Supplemental Materials for "The accumulation of erythrocytes quantified and visualized by Glycophorin C in carotid atherosclerotic plaque reflects intraplaque hemorrhage and pre-procedural neurological symptoms"

Accompanying

### Supplemental Table 1: Confounders included in the model 2 logistic or cox regression models of the association between glycophorin and intraplaque hemorrhage, pre-procedural neurological symptoms and 3-years MACE, and between IPH and MACE, respectively.

| **Glycophorin - IPH** |  |  |
| --- | --- | --- |
|  | Full cohort | Plaque size, hospital of inclusion, history of coronary artery disease |
|  | Male | Plaque size, systolic blood pressure, significant ipsilateral carotid stenosis (50-70%) and critical ipsilateral carotid stenosis (70-99%) |
|  | Female | *None* |
| **Glycophorin - Symptoms** |  |  |
|  | Full cohort | Plaque size, age, gender, hospital of inclusion, smoking status, history of coronary artery disease, history of peripheral arterial occlusive disease, use of antiplatelet drugs and ipsilateral critical carotid stenosis (70-99%) |
|  | Male | Plaque size, age, smoking status, history of peripheral arterial occlusive disease, use of antiplatelet drugs, significant ipsilateral carotid stenosis (50-70%) and critical ipsilateral carotid stenosis (70-99%) |
|  | Female | Plaque size, age, hospital of inclusion and triglyceride levels |
| **Glycophorin - MACE** |  |  |
|  | Full cohort | Plaque size, age, total cholesterol levels, low-density lipoprotein levels, smoking status, history of coronary artery disease, history of peripheral arterial occlusive disease, use of antiplatelet drugs and use of anti-coagulant drugs |
|  | Male | Plaque size, age, smoking status, history of peripheral arterial occlusive disease, use of antiplatelet drugs, use of anti-coagulant drugs, significant ipsilateral carotid stenosis (50-70%) and critical ipsilateral carotid stenosis (70-99%) |
|  | Female | Plaque size, age, hospital of inclusion, total cholesterol and low-density lipoprotein levels |
| **IPH -MACE** |  |  |
|  | Full cohort | History of coronary artery disease |
|  | Male | History of coronary artery disease, diabetes mellitus, history of peripheral interventions and degree of ipsilateral carotid stenosis |
|  | Female | *None* |

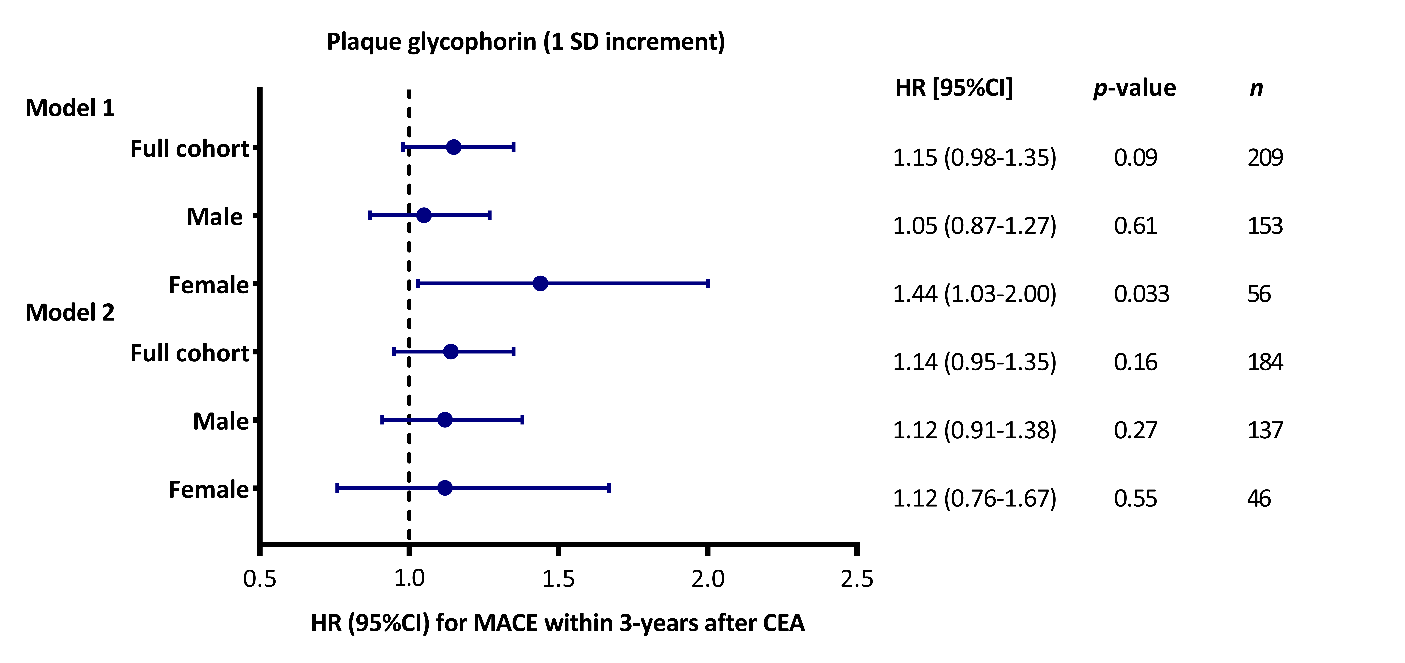

### Supplemental Figure 1: Multivariable associations between plaque glycophorin (1 SD increment) with new major cardiovascular adverse events within 3-years after surgery.

Associations adjusted for plaque size (Model 1) and adjusted for plaque size and confounders (Model 2) are shown. Shown are odd ratios (OR) or hazard ratios (HR) and error bars corresponding to their 95% confidence intervals (CI). The confounders included in the model 2 associations can be found in **Supplemental Table 2**. Multivariable associations of glycophorin C with MACE within 3-years after surgery, as derived from Cox regression analyses.

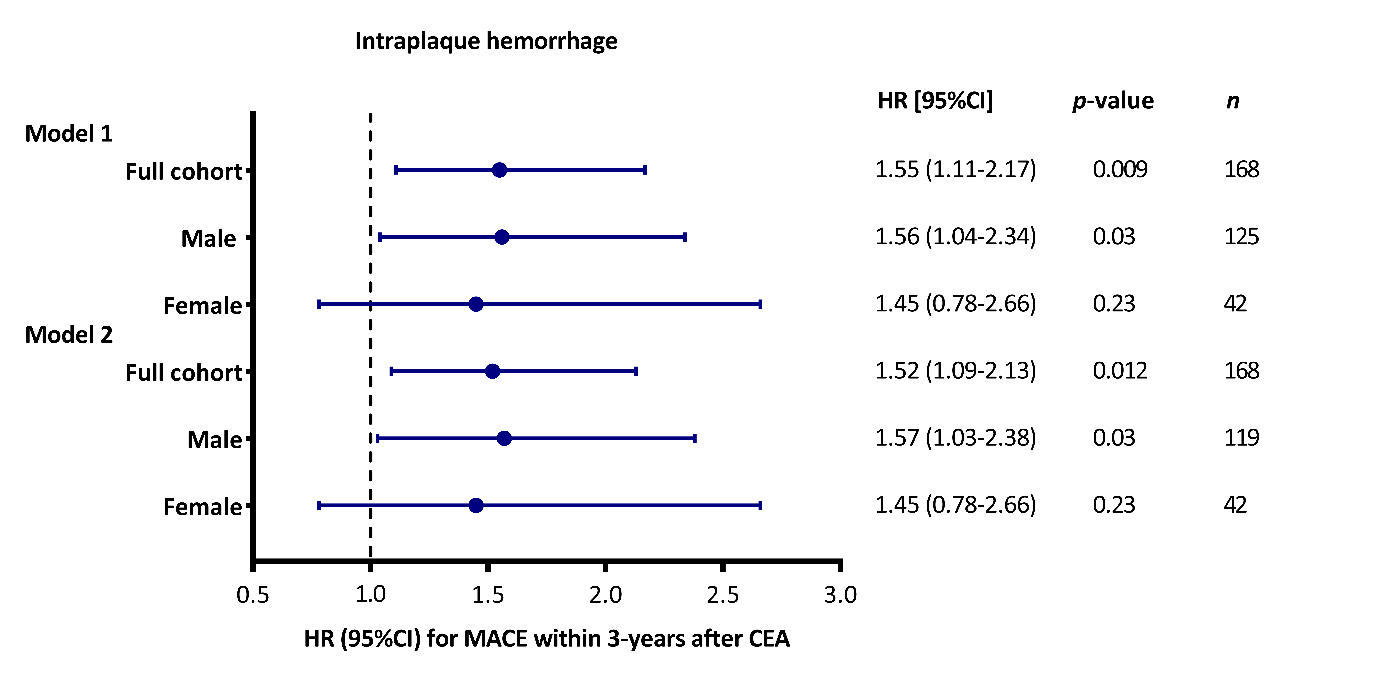

### Supplemental Figure 2: Univariable and multivariable associations between intraplaque hemorrhage with new major cardiovascular adverse events within 3-years after surgery.

Unadjusted associations (Model 1) and associations adjusted for confounders (Model 2) are shown. Shown are hazard ratios (HR) and error bars corresponding to their 95% confidence intervals (CI). The confounders included in the model 2 associations can be found in **Supplemental Table 2**.

### Supplemental Table 2: Descriptive statistics of glycophorin C expressed as percentage glycophorin of total plaque surface described by overall plaque phenotype.

| **Plaque phenotype** | **Mean** | **Max** | **Min** | **Median** | **SE** |
| --- | --- | --- | --- | --- | --- |
| **Fibrous** | 5.532673 | 48.70765 | 0.031439195 | 3.546172 | 0.2778147 |
| **Fibroatheromatous** | 8.440393 | 53.62971 | 0.005309934 | 6.066214 | 0.3750115 |
| **Atheromatous** | 11.780988 | 53.47838 | 0.021295482 | 9.271010 | 0.5014102 |

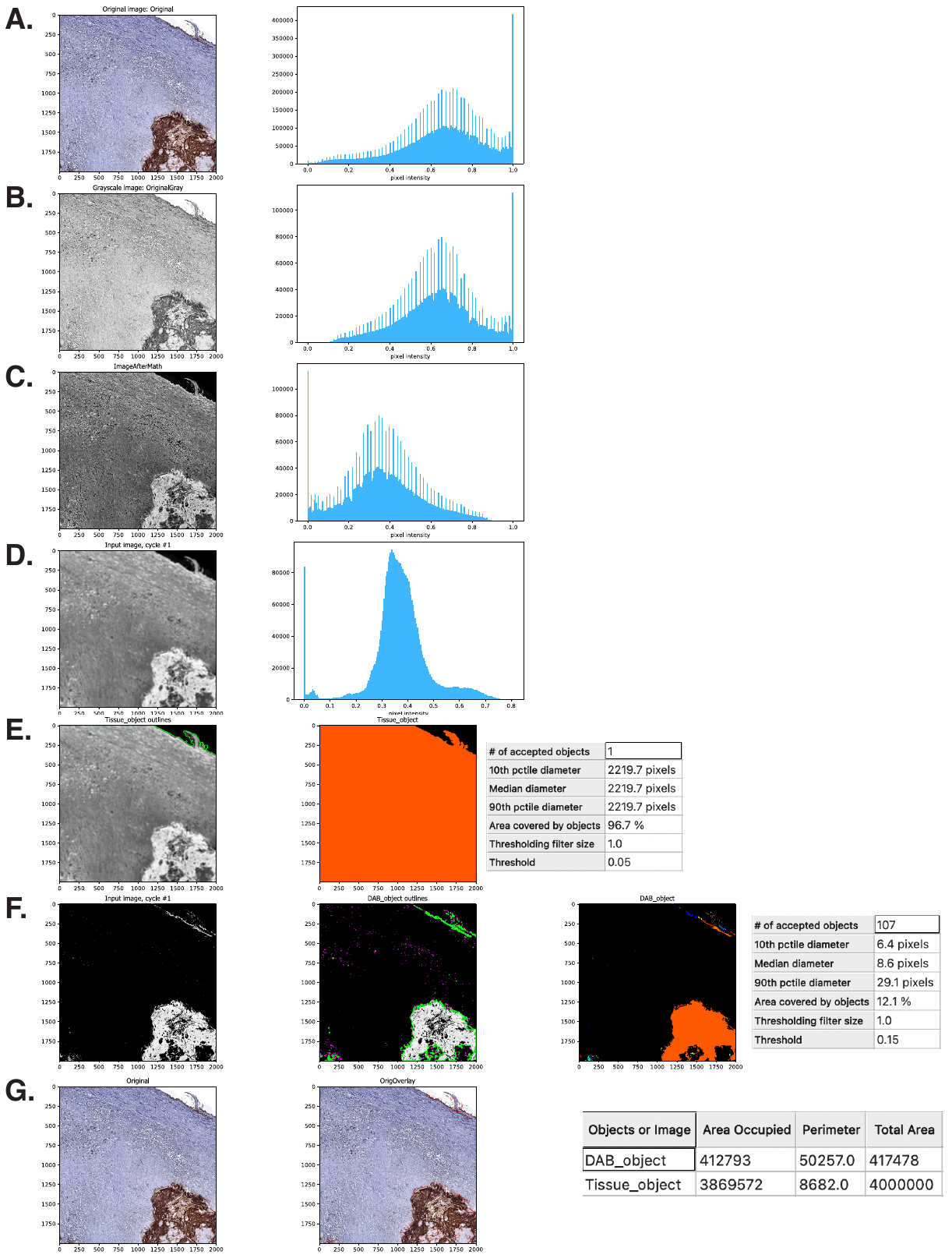

### Supplemental Figure 3: The Glycophorin C CellProfiler pipeline workflow.

**A.** The original image (left) is masked and normalized using slideEntropy and slideNormalize in the slideToolKit workflow and used as input by CellProfiler; the graph (right) shows the tonal distribution in the digital whole-slide image on a RGB scale. **B.** The input image is converted to a gray scaled image (left); the graph (right) shows the tonal distribution in the gray scaled image. **C.** The gray scaled image is inverted, i.e. non-tissue will become black (left); the graph (right) shows the tonal distribution after inverting. **D.** A Gaussian filter is applied to smoothen the image and reduce image artefacts (artifact size 16 pixels) and noise (left); the graph (right) shows the tonal distribution after smoothening. **E.** The tissue area is identified, as demarcated by the green line in the left image; the total tissue area size is calculated in pixels (right image) and tabulated (table). **F.** The stained area is identified, white areas in the left imaged; the stained area is demarcated by a green line in the middle image, areas that are excluded due to size (minimal size 6 pixels) are demarcated in magenta; the total stained area size is calculated in pixels and tabulated (table). **G.** Finally the data for each tile are saved in a comma-separated table, including meta-data such as tile positions, image location, object counts (there could be multiple patches of stained areas or tissue). The original image (left) is used to overlay the stained area (demarcated with red) and the tissue area (demarcated with green).

### Supplemental Table 3: Percentage of participant with missing values.

BMI: body mass index; BP: blood pressure; mmHg: millimeter of mercury; TIA: transient ischemic attack; HDL: high density lipoprotein; LDL: low density lipoprotein; eGFR: estimated Glomerular filtration rate.

|  | **Percentage of participants with missing values before multivariate imputation by chained equations (%)** | **Percentage of participants with missing values after multivariate imputation by chained equations (%)** |
| --- | --- | --- |
| Hospital | 0.0 | 0.0 |
| Age | 0.0 | 0.0 |
| Gender | 0.0 | 0.0 |
| Systolic BP | 11.3 | 11.3 |
| Diastolic BP | 11.3 | 11.3 |
| Hypertension | 1.8 | 1.8 |
| Diabetes mellitus | 0.0 | 0.0 |
| Hypercholesterolemia | 7.3 | 7.3 |
| Total cholesterol | 37.3 | 10.3 |
| HDL | 40.5 | 10.9 |
| LDL | 44.7 | 12.1 |
| Triglycerides | 41.6 | 11.2 |
| BMI | 3.7 | 3.7 |
| eGFR | 3.7 | 1.0 |
| Smoking Status | 4.8 | 4.8 |
| History of coronary artery disease | 0.0 | 0.0 |
| History of peripheral artery disease | 0.0 | 0.0 |
| History of peripheral intervention(s) | 0.3 | 0.3 |
| History of stroke or TIA | 0.0 | 0.0 |
| Use of antihypertensive drugs | 0.2 | 0.2 |
| Use of statins | 0.2 | 0.2 |
| Use of antiplatelet drugs | 0.3 | 0.3 |
| Use of oral anticoagulant drugs | 0.7 | 0.7 |
| Ipsilateral pre-operative stenosis | 2.2 | 2.2 |
| Contralateral stenosis | 8.6 | 3.2 |
